# Genetic and Demographic Determinants of Fuchs Endothelial Corneal Dystrophy Risk and Severity

**DOI:** 10.1101/2024.09.29.24314217

**Authors:** Siyin Liu, Amanda N. Sadan, Nihar Bhattacharyya, Christina Zarouchlioti, Anita Szabo, Marcos Abreu Costa, Nathaniel J. Hafford-Tear, Anne-Marie S. Kladny, Lubica Dudakova, Marc Ciosi, Ismail Moghul, Mark R. Wilkins, Bruce Allan, Pavlina Skalicka, Alison J. Hardcastle, Nikolas Pontikos, Catey Bunce, Darren G. Monckton, Kirithika Muthusamy, Petra Liskova, Stephen J. Tuft, Alice E. Davidson

## Abstract

**Importance:** Understanding the pathogenic mechanisms of Fuchs endothelial corneal dystrophy (FECD) is essential for developing gene-targeted therapies.

**Objective:** To investigate associations between demographic data and age at first keratoplasty in a large genetically refined FECD cohort.

**Design, Setting, and Participants:** This retrospective cohort study recruited 894 individuals with FECD at Moorfields Eye Hospital (London) and General University Hospital (Prague). Ancestry was inferred from genome-wide SNP array data. CTG18.1 status was determined by short tandem repeat and/or triplet-primed PCR. One or more expanded alleles (≥50 repeats) were classified as expansion-positive (Exp+). Expansion-negative (Exp-) cases were whole exome sequenced.

**Main Outcome(s) and Measure(s):** Association between variants in FECD-associated genes, demographic data and age at first keratoplasty.

**Results:** Within the total cohort (n=894), 77.3% were Exp+. The majority of European (668/829, 80.6%) and South Asian (14/22, 63.6%) patients were Exp+, whereas the majority of those with African (30/37, 81.1%), East Asian (2/3,66.7%), and American Admixture (2/3, 66.7%) ancestry were Exp-. The percentage of females was significantly higher (151, 74.4%) in the Exp-cohort compared to the Exp+ (395, 57.2%; *P*<.001). The median (IQR) age at first keratoplasty of the Exp+ patients (68.2 [63.2–73.6] years) was older than the Exp-patients (61.3 [52.6–70.4] years; *P*<.001). The CTG18.1 repeat length of the largest expanded allele within the Exp+ group was inversely correlated with the age at first keratoplasty (r = -0.087 [95% CI: -0.162 to -0.012], *P*=.02). The ratio of biallelic to monoallelic expanded alleles was higher in the FECD cohort compared to an unaffected control group (*P*<.001), indicating that two Exp+ alleles increase disease penetrance compared to one expansion. We only identified potentially pathogenic variants (MAF <0.01; CADD >15) in FECD-associated genes in 13 (10.1%) Exp-individuals.

**Conclusions and Relevance:** CTG18.1 expansions are present in most European and South Asian patients with FECD. Repeat length and zygosity status of CTG18.1 modify disease severity and penetrance. Known disease-associated genes account for only a minority of Exp-cases, with unknown risk factors determining disease in the rest of this subgroup. These data have important implications for the development of future FECD gene-targeted therapies.

**Key Points:** *Question:* How do demographics and genetic risk factors determine FECD disease risk and severity?

*Findings:* This multi-center FECD cohort (n=894) study reveals *TCF4* repeat expansions (CTG18.1) underlie disease in 77.3% of total cases, with longer repeats and biallelic expansion correlating with earlier keratoplasty and increased penetrance, respectively. Female overrepresentation is driven by cases without CTG18.1 expansions, where missing heritability remains high.

*Meaning:* Demographic factors and molecular diagnosis, including CTG18.1 repeat length and expanded allele dosage, are clinically relevant metrics that should inform future therapeutic strategies and clinical trial design.

## Introduction

Fuchs endothelial corneal dystrophy (FECD) is a bilateral, progressive disease of the corneal endothelium that is a leading indication for keratoplasty in high-income countries.^1,2^ It is a genetically heterogeneous, variably penetrant, autosomal dominant trait. Most studies report a preponderance of females, with a ratio of 1.5 to 3.7 female per male.^3–9^ The disease appears to be more prevalent in European than East Asian or Middle Eastern populations.^10–14^ A recent FECD comorbidity association study demonstrated that female sex and European ancestry increase the risk of developing FECD by 4.6 fold and 5.5 fold, respectively.^15^ Depending on ancestry, 17% to 81% of FECD patients in these cohorts have one or more expanded copies of an intronic CTG repeat within the *TCF4* gene (termed CTG18.1; MIM: *602272.0007),^6–9,16–25^ making it, by far, the most common trinucleotide repeat expansion disease. Other rarer genetic causes have been identified through linkage analysis and candidate gene screening within familial cohorts.^26,27^ For example, heterozygous missense variants in *COL8A2* cause an early-onset and phenotypically distinct form of the disease.^28,29^ Rare variants in other genes, including *SLC4A11, ZEB1, AGBL1, LOXHD1* and *TCF4,* have also been associated with FECD, though several findings have not been replicated.^13,26,30–33^ In addition, three FECD genome-wide association studies (GWAS) have collectively identified twelve significant genomic loci but, excluding CTG18.1 expansion status, the causal risk variants driving these association signals remain elusive.^34–36^

Gene-targeted interventions to prevent or delay FECD progression are in development.^37–41^ However, their success will rely on identifying at-risk individuals before corneal endothelial function deteriorates and sight loss occurs. Here, we present an in-depth analysis of a large and extensively genotyped FECD cohort, providing novel clinical, demographic and genetic insights.

## Methods

### Subject recruitment

We recruited participants at Moorfields Eye Hospital (MEH), London and the General University Hospital(GUH), Prague, from September 2009 to July 2023, following the tenets of the Declaration of Helsinki. The study was approved by the Research Ethics Committees of University College London (UCL) (22/EE/0090), Moorfields Eye Hospital (MEH) London (13/LO/1084), or the General University Hospital (GUH) Prague (151/11 S-IV). All participants were diagnosed with FECD based on the documented finding of confluent corneal guttae seen by slit-lamp examination. They provided written consent and whole blood or saliva for DNA extraction. The potential effect of CTG18.1 genotype on phenotypic outcome was evaluated using two clinical parameters: A) age at recruitment (at date of whole blood or saliva sample collection); B) the age at first keratoplasty (at date of their first keratoplasty), which could be before or after recruitment. To prevent the confounding effect of traumatic endothelial cell loss, patients with a history of intraocular surgery, including cataract extraction, were excluded. Similarly, we only included patients who had a primary endothelial keratoplasty with or without phacoemulsification. The study workflow is summarized in **eFigure 1**.

**Figure 1.**
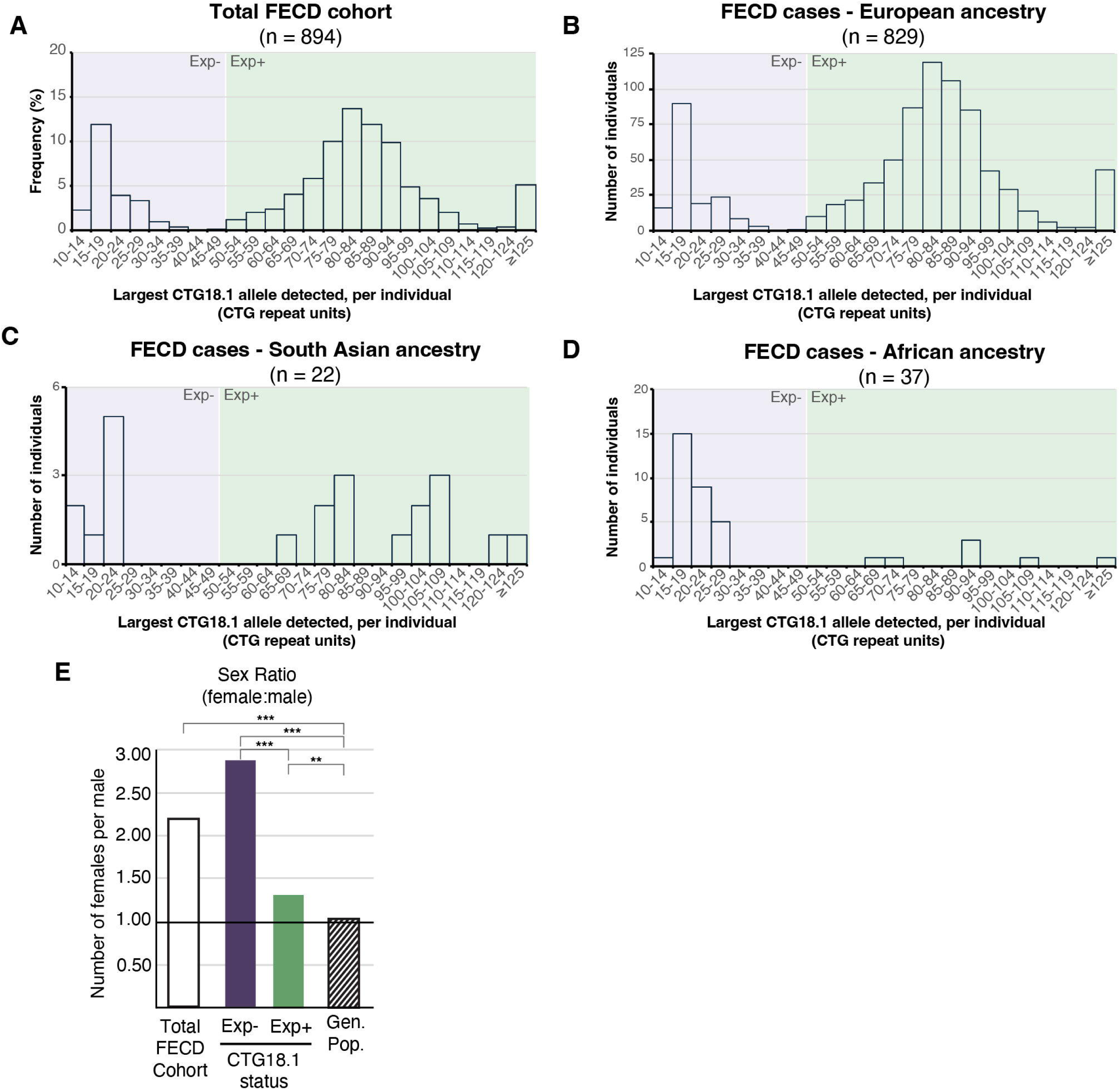
CTG18.1 repeat length distributions vary with ancestry and sex within a large FECD patient cohort. Frequency histograms comparing the relative distribution of CTG18.1 repeat length within the (**A**) total cohort (894), and the allelic distributions of CTG18.1 repeat length within (**B**) European (EUR; 829), (**C**) South Asian (SAS; 22) and (**D**) African (AFR; 37) ancestry groups. The percentage of European patients with a CTG18.1 expansion (80.7%) was greater than that of non-Europeans (35.4%). A larger proportion of South Asian patients also harbored at least one expanded CTG18.1 allele (63%), compared to patients of African ancestry (18%). (**E**) Bar chart of the sex ratio across the total cohort (2.20 female:male, black bar) and subgroups stratified by CTG18.1 status: Exp-group (2.88 female:male, purple bar), Exp+ group (1.34 female:male, green bar), General population over 40 in England and Wales (1.04 female:male, striped bar). Abbreviation: CTG18.1 expansion positive allele defined as ≥50 CTG repeats; Exp-, CTG18.1 expansion negative allele defined as <50 CTG repeats.

### DNA extraction and CTG18.1 genotyping

Genomic DNA was extracted from whole blood or saliva using a Gentra Puregene Blood kit (Qiagen) or Oragene saliva kit (Oragene OG-300, DNA Genotek). All samples were analyzed using a previously-described short tandem repeat (STR)-polymerase chain reaction (PCR) assay.^16,18,42^ Triplet repeat-primed (TP)-PCR was subsequently performed if only one CTG18.1 allele was detected to determine if an allele longer than the STR-PCR detection maximum (∼125 repeats) was present. We defined cases with one or both alleles having ≥50 repeats as expansion-positive (Exp+) and those with biallelic alleles of <50 repeats as expansion-negative (Exp-).^17,42^

### Ancestry and relatedness

We genotyped all participants using a UK Biobank Axiom Array (Applied Biosystems). Genotypes were called using Axiom Analysis Suite software. Ancestry was inferred by principal component analysis (PCA) (FRAPOSA).^43^ We used 2,492 unrelated samples with known ancestry from the 1000 Genomes Project as a reference panel. Kinship analysis was performed using KING.^44^ Probands were defined as the first recruited individual within such kinships, and all cases identified as 2nd-degree cousins or more closely related to probands were excluded (kinship coefficient > 0.0884).

### Exome sequencing and rare variant analysis pipeline

Exome libraries were generated using a SureSelect Human All Exome V6 capture kit (Agilent) or a SeqCap EZ MedExome Enrichment Kit (Roche) and sequenced on either a HiSeq 4000 or 2500 platform (Illumina). Raw sequencing data were aligned using Burrows-Wheeler Aligner (BWA, 0.7.17).^45,46^ Variants and indels were called according to the Genome Analysis Toolkit Haplotypecaller (GATK, v4.4).^47^ Aligned data were interrogated for rare and potentially disease-associated coding variants in previously implicated in FECD genes: *COL8A2*, *ZEB1*, *SLC4A11*, *AGBL1*, *LOXHD1* and *TCF4.*^26,27,33^ Variants were annotated using Ensembl VEP (106.1),^48^ with the Combined Annotation Dependent Depletion (CADD, v1.6)^49^ and REVEL^50^ plugins. We defined variants of interest as having a CADD score >15 and a minor allele frequency (MAF) <0.01 in the Genome Aggregation Database (gnomAD, v3.1.2) in all genetic ancestry groups, excluding the Amish population.^51^ Variants of interest were verified by Sanger sequencing. SpliceAI was used to assess the effect of splice region variants on splicing.^52^

### Corneal endothelial transcriptome analysis

Cultured corneal endothelial cell (CEC) transcriptomes from four healthy control adults were queried to determine the relative abundance of the genes expressed within the corneal endothelium (EGAS50000000303).^33^ Briefly, FASTQ files were quantified with Salmon (GRCh38.p13, Ensembl v100, V1.4.0)^53^ and tximport (v.1.30.0)^54^ to generate normalized TPM gene-level counts. Genes with expression levels (TPM) ≤ 0.02 were considered not expressed within the corneal endothelium.

### Statistical analysis

Statistical analysis was performed using R (version 4.0.2, R Foundation for Statistical Computing). Data normality was assessed using Kolmogorov–Smirnov/Shapiro–Wilk tests. We used the χ2 test to compare categorical data. Wilcoxon signed-rank test was used to analyze non-parametric continuous variables. A linear regression model assessed the association between CTG18.1 repeat length of the largest expanded allele and age at first keratoplasty. Cases with repeat lengths of ≥125 repeats, which exceed the detection limit of STR-PCR, and cases with biallelic Exp+ where the repeat lengths of either allele could not be determined by the STR-PCR assay, were excluded from the regression analysis. Allele frequencies of FECD cases and unaffected, aged controls, previously reported,^17^ were calculated to derive the observed and expected monoallelic to biallelic Exp+ allelic ratio, respectively. The observed and expected ratios of monoallelic to biallelic Exp+ cases were compared. *P* valuesL<.05 were classified as significant.

## Results

### FECD cohort sex and ancestry vary with CTG18.1 allelic distributions

We recruited 918 patients with FECD. Twenty-four were determined to be closely related and excluded, leaving 894 probands, of which 546 (61.1%) were females. PCA of genome-wide SNP array data showed that 829 (92.7%) were European (**Table 1**). CTG18.1 genotyping revealed that 691 (77.3%) participants had at least one expanded copy of the CTG18.1 allele, and 46 (5.1%) had bi-allelic expansions. More European patients had a CTG18.1 expansion (668, 80.6%) compared to non-Europeans (23, 35.4%, *P*<.001), in agreement with gnomAD (v3.1.2), which shows Europeans have the highest population frequency of CTG18.1 expansion.^55^ By comparing these data to the age and ethnicity-matched control cohort,^17^ harboring at least one expanded CTG18.1 allele conferred >78- fold risk for FECD in patients of European ancestry (odd ratio [OR] = 78.5; 95% CI: 50.3 to 122.6, *P*<.001). Overall, there was a significantly lower proportion of Exp+ African cases (7, 18.9%) compared to European cases (*P*<.001), while the proportion of Exp+ cases between South Asian (14, 63.6%) and European groups was similar (*P*=0.09; **Table 1**; **Figure 1A**).

**Table 1.** Summary of Fuchs endothelial corneal dystrophy (FECD) patient cohort demographics and CTG18.1 expansion status. Exp+, CTG18.1 expansion positive cases defined as one or both expanded alleles (≥50 CTG repeats); Exp-, CTG18.1 expansion negative cases defined as biallelic alleles of <50 repeats; MEH, Moorfields Eye Hospital; GUH, General University Hospital. The cohort data presented has been updated from an earlier publication.^17^ ^a^Determined by principal component analysis using FRAPOSA.^43^

|  | Number of cases | CTG18.1 Exp+ |  |  | CTG18.1 Exp- |
| --- | --- | --- | --- | --- | --- |
| | | Cases with $\geq 1$ expanded allele | Monoallelic expanded cases | Biallelic expanded cases | |
| **Genotyped patients** | 894 | 691 (77.3%) | 645 (72.1%) | 46 (5.1%) | 203 (22.7%) |
| **Sex** |  |  |  |  |  |
| Females | 546/894 (61.1%) | 395/691 (57.2%) | 371/645 (57.5%) | 24/46 (52.2%) | 151/203 (74.4%) |
| Males | 348/894 (38.9%) | 296/691 (42.8%) | 274/645 (42.5%) | 22/46 (47.8%) | 52/203 (25.6%) |
| **Ancestry<sup>a</sup>** |  |  |  |  |  |
| European | 829 (92.7%) | 668/829 (80.6%) | 622/829 (75.0%) | 46/829 (5.5%) | 161/829 (19.4%) |
| African | 37/894 (4.1%) | 7/37 (18.9%) | 7/37 (18.9%) | 0/37 (0.0%) | 30/37 (81.1%) |
| South Asian | 22/894 (2.5%) | 14/22 (63.6%) | 14/22 (63.6%) | 0/22 (0.0%) | 8/22 (36.4%) |
| East Asian | 3/894 (0.3%) | 1/3 (33.3%) | 1/3 (33.3%) | 0/3 (0.0%) | 2/3 (66.7%) |
| American Admixture | 3/894 (0.3%) | 1/3 (33.3%) | 1/3 (33.3%) | 0/3 (0.0%) | 2/3 (66.7%) |
| **Recruitment Site** |  |  |  |  |  |
| MEH | 563 | 430/563 (76.4%) | 401/563 (71.2%) | 29/563 (5.2%) | 133/563 (23.6%) |
| GUH | 331 | 262/331 (79.2%) | 245/331 (74.0%) | 17/331 (5.1%) | 69/331 (20.8%) |

The high proportion of females in the total FECD cohort (2.20 female-to-male ratio) validates numerous previous reports (**Table 1**). For comparison, females comprise 52.0% (1.04 ratio) of the population of England and Wales over 40 years of age,^56^ which is significantly lower than the sex ratio in the total FECD cohort (P<.001). The Exp+ subgroup also exhibited a higher proportion of females (1.34 ratio) than the general population of England and Wales (*P*=.007). In the Exp- subgroup 74.4% were female (2.88 ratio), which is significantly higher than both the general population (*P*<.001) and the Exp+ subgroup (*P*<.001; **Figure 1B**).

### CTG18.1 repeat length and expanded allele dosage modify the age at first keratoplasty and disease penetrance, respectively

The median (interquartile range [IQR]) age at recruitment of the total FECD cohort was 69.7y (62.7–76.1), though the Exp- patients (66.7y [53.7–74.7]) were significantly younger than Exp+ patients (70.2y [64.7–76.4], *P*<.001; **Table 2**). Whilst both male and female Exp+ patients were recruited at a similar age, Exp- males tended to be recruited at a younger age than females (62.6y [50.8–73.6] vs 67.7y [55.8–75.1], *P*=.13). After excluding cases that did not meet the inclusion criteria for genotype-keratoplasty data analysis, a higher proportion of Exp+ patients (382, 58.9%) had keratoplasty than the Exp- group (59, 34.9%; **eTable 1**).

**Table 2.**
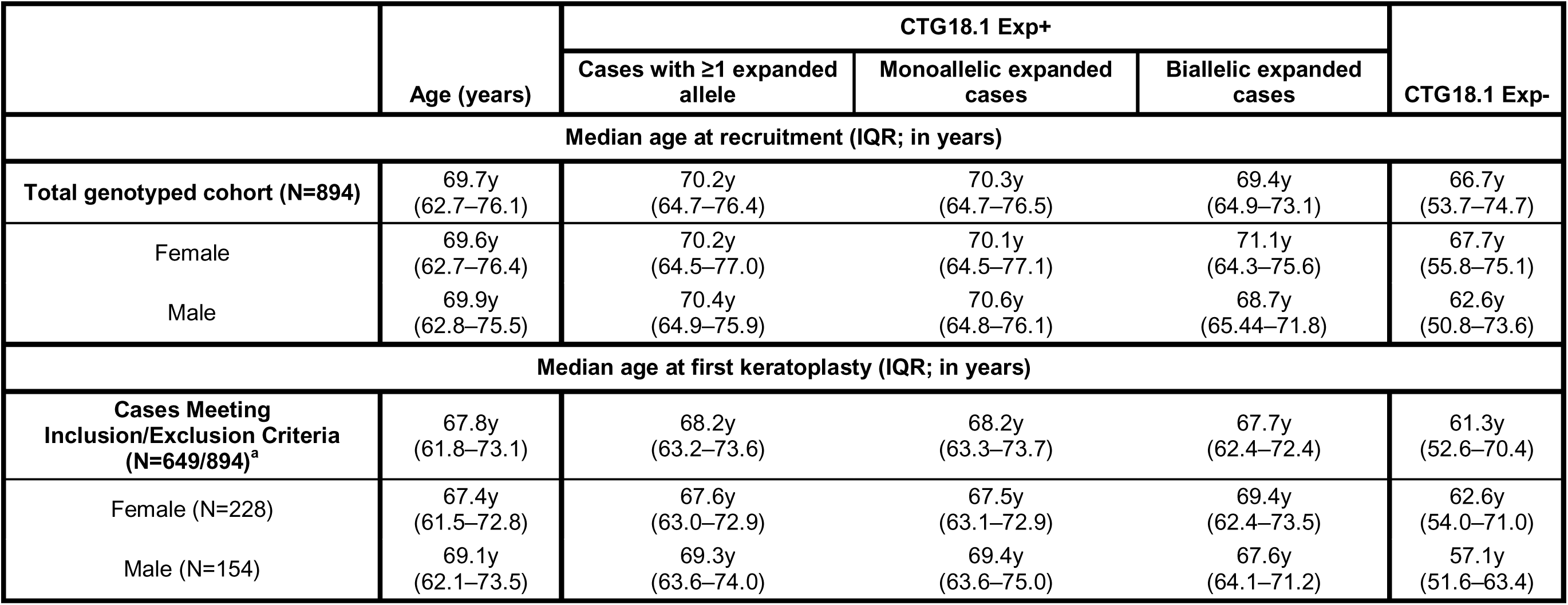
Age at recruitment and first keratoplasty of Fuchs endothelial corneal dystrophy (FECD) patient cohort stratified by sex and CTG18.1 genotype. Exp+, CTG18.1 expansion positive allele defined as ≥50 CTG repeats; Exp-, CTG18.1 expansion positive allele defined as <50 CTG repeats; IQR, interquartile range. ^a^Inclusion criteria: endothelial keratoplasty, whether as a standalone procedure, combined with phacoemulsification, or sequentially planned after phacoemulsification; Exclusion criteria: Penetrating keratoplasty, prior intraocular surgery, and unspecified keratoplasty type; the denominators include the total cohort, encompassing both operated and unoperated cases, after excluding those that meet the exclusion criteria.

|  | Age (years) | CTG18.1 Exp+ |  |  | CTG18.1 Exp- |
| --- | --- | --- | --- | --- | --- |
|  |  | Cases with ≥1 expanded allele | Monoallelic expanded cases | Biallelic expanded cases |  |
| Median age at recruitment (IQR; in years) |  |  |  |  |  |
| Total genotyped cohort (N=894) | 69.7y<br>(62.7–76.1) | 70.2y<br>(64.7–76.4) | 70.3y<br>(64.7–76.5) | 69.4y<br>(64.9–73.1) | 66.7y<br>(53.7–74.7) |
| Female | 69.6y<br>(62.7–76.4) | 70.2y<br>(64.5–77.0) | 70.1y<br>(64.5–77.1) | 71.1y<br>(64.3–75.6) | 67.7y<br>(55.8–75.1) |
| Male | 69.9y<br>(62.8–75.5) | 70.4y<br>(64.9–75.9) | 70.6y<br>(64.8–76.1) | 68.7y<br>(65.44–71.8) | 62.6y<br>(50.8–73.6) |
| Median age at first keratoplasty (IQR; in years) |  |  |  |  |  |
| Cases Meeting Inclusion/Exclusion Criteria (N=649/894)<sup>a</sup> | 67.8y<br>(61.8–73.1) | 68.2y<br>(63.2–73.6) | 68.2y<br>(63.3–73.7) | 67.7y<br>(62.4–72.4) | 61.3y<br>(52.6–70.4) |
| Female (N=228) | 67.4y<br>(61.5–72.8) | 67.6y<br>(63.0–72.9) | 67.5y<br>(63.1–72.9) | 69.4y<br>(62.4–73.5) | 62.6y<br>(54.0–71.0) |
| Male (N=154) | 69.1y<br>(62.1–73.5) | 69.3y<br>(63.6–74.0) | 69.4y<br>(63.6–75.0) | 67.6y<br>(64.1–71.2) | 57.1y<br>(51.6–63.4) |

The median age at first keratoplasty for the Exp+ patients (68.2y [63.2–73.6]) was significantly older than for the Exp- patients (61.3y [52.6–70.4], *P*<.001; **Table 2**). This is likely due to a broader age distribution within Exp-, where there was a subset who had surgery at a relatively young age (Exp+ 40y–95y vs Exp- 22y–87y, *P*<.001) (**Figure 2A**).

**Figure 2.**
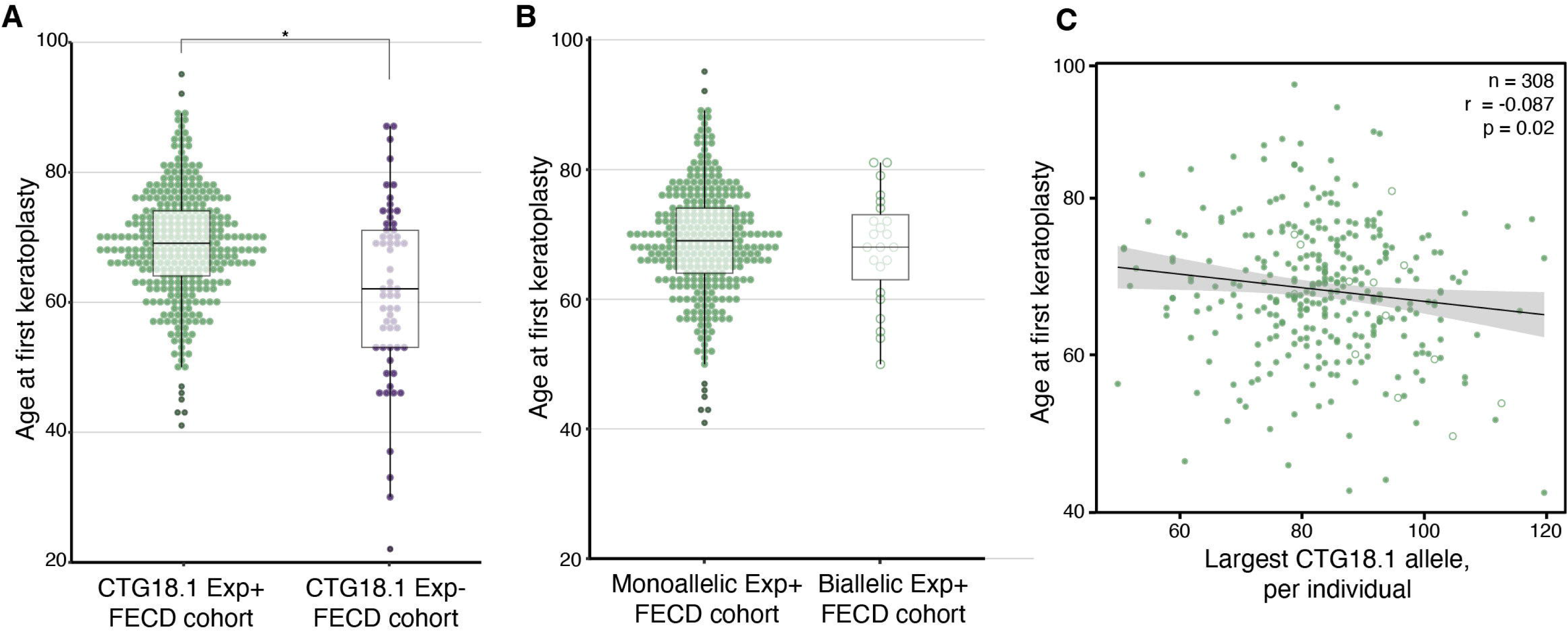
Age at first keratoplasty varies depending on CTG18.1 expansion status, repeat length and zygosity. (**A**) Age at first keratoplasty (median [IQR]) was more heterogeneous in the expansion-negative (Exp-) group (61.3y [52.6–70.4]) compared to the expansion-positive (Exp+) group (68.2y [63.2–73.6]), including both mono- and bi-allelic Exp+ cases (*P*<.001). (**B**)The median age at first keratoplasty in FECD patients with biallelic Exp+ (67.7y [62.4–72.4]) was not was not statistically significantly different (*P*<.69) than in monoallelic Exp+ patients (68.2y [63.3–73.7]).(**C**) The scatter plot demonstrates a negative correlation between the CTG18.1 repeat length and age at first keratoplasty in Exp+ patients (r = -0.087 [95% CI: -0.162 to -0.012], P = .02). *Green dots*, monoallelic Exp+ cases; *green open circles,* biallelic Exp+ cases.

Although the median age at first keratoplasty was lower for patients with biallelic CTG18.1 Exp+ (67.7y [62.4–72.4]) compared to those with monoallelic expansions, the difference did not reach significance (68.2y [63.3–73.7], *P<.69*; **Figure 2B**; **Table 2**). Notably, the observed ratio of biallelic to monoallelic Exp+ cases, derived from homozygous to heterozygous Exp+ allelic ratios, was significantly higher in the FECD cohort (1:14) compared to the expected ratio of Exp+ cases in an aged, unaffected group^17^ (1:94) (expected vs observed biallelic Exp+ cases: 7 vs 46; *P<.001*; **eTable 2**), suggesting that disease penetrance is higher in carriers of two expanded copies of CTG18.1.

Within the refined Exp+ patient group with sized CTG18.1 alleles (308/382; **eTable 3**), linear regression demonstrated a significant negative correlation between the CTG18.1 repeat length of the single largest expanded allele and age at first keratoplasty (r = -0.087 [95% CI: -0.162 to -0.012], *P*=.02; **Figure 2C)**.

### Rare coding variants in FECD-associated genes account for a minor fraction of missing heritability in Exp- cases

To explore the missing heritability in the Exp- group, exome data was generated for 128 Exp- patients. FECD-associated genes were interrogated for rare and potentially deleterious variants in conjunction with bulk CEC-specific RNAseq data. Analysis of the transcriptomic data revealed that neither *LOXHD1* nor *AGBL1* are expressed (**eTable 4**). This finding, in conjunction with the fact that neither gene has been replicated as FECD-associated,^26^ led us to discount variants in these genes. Within the remaining robustly validated gene set (*COL8A2*, *SLC4A11*, *ZEB1,* and *TCF4*), we only identified potentially disease-associated variants in 13 (10.1%) of 128 patients (**Table 3**).

**Table 3.**
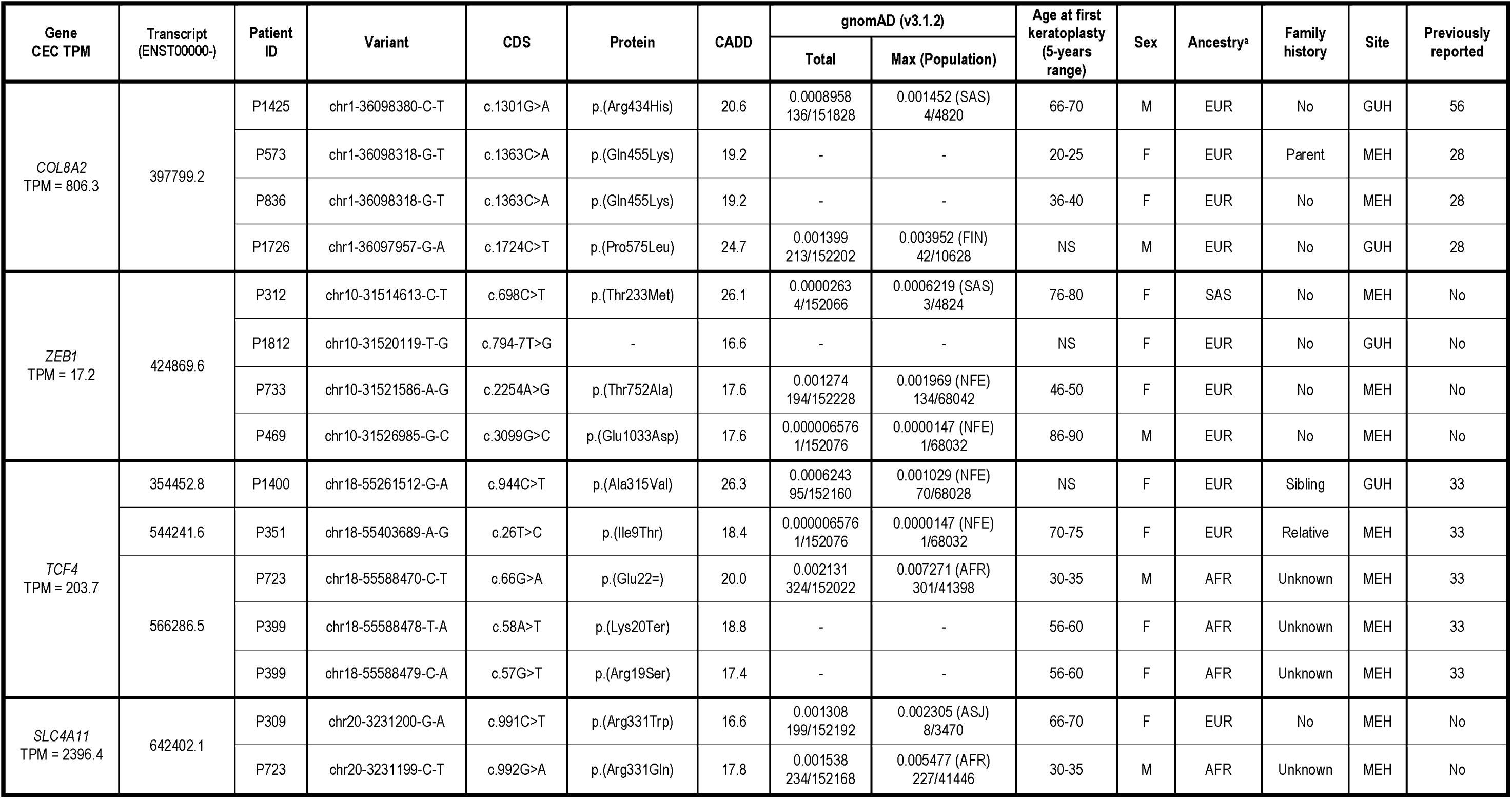
Summary of rare and potentially pathogenic variants identified in FECD-associated genes from 128 FECD CTG18.1 Exp- probands analysed by exome sequencing. Relatives were not included in the main analysis even though identified here. The identifiers used in this study do not disclose the identity of any participants beyond the immediate research team. CEC, corneal endothelial cells; TPM, transcript per million; CADD, Combined Annotation Dependent Depletion; MAF, minor allele frequency; EUR, European; AFR, African American/African; FIN, Finnish; NFE, non-Finnish European, ASJ, Ashkenazi Jews; M, male; F, female; NS, no surgery; MEH, Moorfields Eye Hospital London; GUH, General University Hospital Prague. ^a^FRAPOSA predicted ancestry.

Four Exp- patients had three qualifying heterozygous *COL8A2* missense variants (**Table 3**). Two harbored the same pathogenic missense variant, c.1363C>A p.(Gln455Lys), previously established to cause early-onset FECD (MIM #136800).^28^ Notably, both cases had corneal transplantation in their second or third decade (**Table 3**). Patients P1425 and P1726 harbored p.(Arg434His) and p.(Pro575Leu) variants, but without early-onset disease. P1425 had a keratoplasty in late 60s, whilst P1726 was recruited in his early 60s but had not undergone surgery. Both variants have been associated with FECD, although lack of segregation with disease has been reported independently, suggesting they may be associated with incomplete penetrance or are non-causal.^28,57^

Two cases, P309 and P723, harbored heterozygous qualifying *SLC4A11* missense variants not previously associated with FECD. Interestingly, both variants alter the same amino acid residue: p.(Arg331Trp) and p.(Arg331Gln) (**Table 3**). In four cases, we identified qualifying *ZEB1* variants, including three heterozygous missense variants p.(Thr233Met), p.(Thr752Ala) and p.(Glu1033Asp) and a heterozygous splice region variant, c.794-7T>G, not predicted by SpliceAI to impact the splicing of any *ZEB1* transcripts (acceptor loss score Δ0.04) (**Table 3**).

There were five qualifying *TCF4* variants in four previously reported patients,^33^ including three missense and two potentially loss-of-function variants (**Table 3**). In case P399, two consecutive missense and nonsense variants occurred *in cis*; c.[57G>T;58A>T]; p.[(Arg19Ser;Lys20*)]. For P723, the rare variant resulted in a synonymous change, c.66G>A p.(Glu22=), that is predicted to result in the loss of a native donor site (SpliceAI donor loss score Δ0.78).^33^

## Discussion

Here we present the largest comprehensively genetically interrogated cohort with FECD reported to date, and show for the first time that: i) dosage of expanded CTG18.1 alleles modifies penetrance, ii) variants in known associated genes only account for a minority of missing heritability in Exp- cases, iii) and the preponderance of female disease is largely driven by Exp- cases.

The prevalence of CTG18.1 expansions varies between ethnic groups, with a reported allele frequency of 2.95% in European, and 0.7%–1.8% in non-European populations.^55^ Here we also confirm our previous finding^17^ that a single expanded allele confers ≥78-fold increased risk of developing FECD. Thus the reported higher prevalence of FECD may be explained by the higher frequency of CTG18.1 expansions in the European population.^10–14^ We found that the proportion of South Asian patients (63.6%) with a CTG18.1 expansion was higher than in previous reports (17.3%-34.0%)^6,21^ which suggests that CTG18.1 expansion may also be a common driver of FECD in this population.

Studies have used a range of clinical metrics to examine the effect of CTG18.1 expansions on disease severity.^19,23,24,58^ In the majority of these studies, the history of cataract extraction and the type of keratoplasty performed were not considered. Our study aimed to examine the inherent biological link between genotype and phenotype, whilst making our best effort to control for potential external influences that could distort the relationship. Hence, strict exclusion criteria were applied for our genotype-phenotype analyses. Our data demonstrate that the length of the largest expanded allele inversely correlates with the age at first keratoplasty. However, the correlation was modest, suggesting that age at first keratoplasty may be too crude a surrogate marker of disease severity and that other genetic or environmental factors modify the phenotype. Future longitudinal studies involving early screening with genotyping will likely improve our understanding of the impact of CTG18.1 repeat length on FECD onset and progression. Repeat length is established as a predictor of age at onset in some repeat-mediated diseases,^59,60^ but the correlation is weak or absent in others.^61–65^ It is also possible that there may be a maximum repeat length threshold above which the phenotypic effect is constant.

In this study, DNA from blood/saliva was used to estimate the inherited allele length. We have previously shown that individuals with ≥50 CTG18.1 repeats detected in blood/saliva consistently display molecular hallmarks of repeat-mediated pathology in their CECs.^33,42^ However, it is important to recognise that expanded CTG18.1 alleles are consistently much larger in affected CECs due to somatic instability.^42^ Nonetheless, the inherited allele length estimates from stable cell populations (i.e. blood/saliva) are considered informative for genotype-phenotype correlations, as shown in previous studies of repeat-mediated disease.^66^

We observed a strong, approximately sevenfold, enrichment of biallelic expansion cases in our cohort, suggesting that two copies of the expanded repeat increase disease penetrance. However, patients with a biallelic CTG18.1 expansion did not have a younger age at first keratoplasty compared to those with a monoallelic expansion. Soliman et al. also found no differences in severity between these two groups when they compared clinical metrics such as Krachmer grade, central corneal thickness, and the proportion who had a keratoplasty.^23^ Thus, two copies of the expanded repeat appear to increase disease penetrance without resulting in detectable signs of increased disease severity in the patient population.

Our data demonstrated that the Exp- group is more ethnically and phenotypically diverse. Despite 10% (13/128) of the exome-sequenced Exp- cases harboring rare qualifying variants in previously reported FECD genes, only one *COL8A2* variant (p.Gln455Lys) has previously been robustly demonstrated as an established cause of FECD.^26,28,67^ Additional analysis is required to validate all remaining variants reported here. Furthermore, future in-depth genomic interrogation will be required to identify other rare Mendelian causes and/or complex genetic risk factors of disease that may in-part explain the missing heritability in the Exp- subgroup, though we cannot exclude the possibility that some cases with the FECD phenotype will not have a genetic basis for their disease. The high female preponderance in the Exp- subgroup highlights a role for sex-specific factors underlying FECD in some cases. For example, dysregulation of estrogen metabolite pathways in FECD CECs and sex-specific sensitivity to UV-induced mitochondrial damage *in vitro* and in animal models have been reported.^68–71^ Thus it is plausible that behavioral^72^ and biological sex differences play a more critical role in FECD cases in the absence of established genetic causes or risk factors.

## Limitations

The majority of patients included in this study are European, which should be noted when contextualizing conclusions regarding ancestry. Accurately determining the age of onset in FECD was impossible as the disease can be asymptomatic for many years. We, therefore, used two surrogates to estimate disease severity: the age at recruitment and the age at first keratoplasty in either eye. Both of these parameters are likely to be affected by uncontrolled variables such as referral practice and patient preference.

## Conclusions

Comprehensive genetic interrogation of this multi-centre FECD cohort provides novel insight into this heterogeneous disease, such as the effect of genotype on phenotypic outcomes.

However, a significant proportion of cases remain genetically unsolved. Several novel CTG18.1-targeted interventions are in development, which may reduce the demand for corneal donor tissue.^17,37,38,40,41,73,74^ Our data indicates that CTG18.1 zygosity status and repeat length of the expanded allele should be included in the design of clinical trials. The success of any of these approaches depends upon population screening to identify individuals with CTG18.1 expansions before irreversible damage occurs. The higher prevalence of CTG18.1 expansions among European and South Asian patients means these populations are particularly well-positioned to benefit from the development of CTG18.1- targeted therapies once integrated into clinical practices.

## Supporting information

eFigure 1

eTable 1

eTable 2

eTable 3

eTable 4

## Data Availability

In this study we used patient identifiable information. For this reason raw data can not be made publicly available due to confidentiality considerations.

## Acknowledgements

Jana Jedlickova and Beverly Scott for technical support. The authors declare no competing interests. AED has previously acted as a paid consultant for Triplet Therapeutics Ltd, LoQus23 Therapeutics Ltd, Design Therapeutics Ltd and had a research collaboration with ProQR Therapeutics. AED has an ongoing research collaboration with Prime Medicine. This work was performed within the framework of ERN-EYE.

## Author contributions

**Concept and design:** Liu, Sadan, Bhattacharyya, Zarouchlioti, Monckton, Muthusamy, Liskova, Tuft, Davidson

**Data acquisition and analysis:** Liu, Sadan, Bhattacharyya, Zarouchlioti, Szabo, Costa, Hafford-Tear, Kladny, Dudakova, Ciosi, Moghul, Wilkins, Allan, Skalicka, Bunce, Muthusamy, Liskova, Tuft, Davidson

**Drafting of the manuscript:** Liu, Bhattacharyya, Zarouchlioti, Costa, Liskova, Tuft, Davidson

**Critical review of the manuscript for important intellectual content:** All authors

**Statistical analysis:** Liu, Bhattacharyya, Zarouchlioti, Ciosi, Bunce, Monckton

**Obtained funding:** Liu, Liskova, Tuft, Davidson

**Supervision:** Monckton, Hardcastle, Pontikos, Muthusamy, Liskova, Tuft, Davidson

