## Supplementary figures and images for "Genetic and Demographic Determinants of Fuchs Endothelial Corneal Dystrophy Risk and Severity"

### eFigure 1

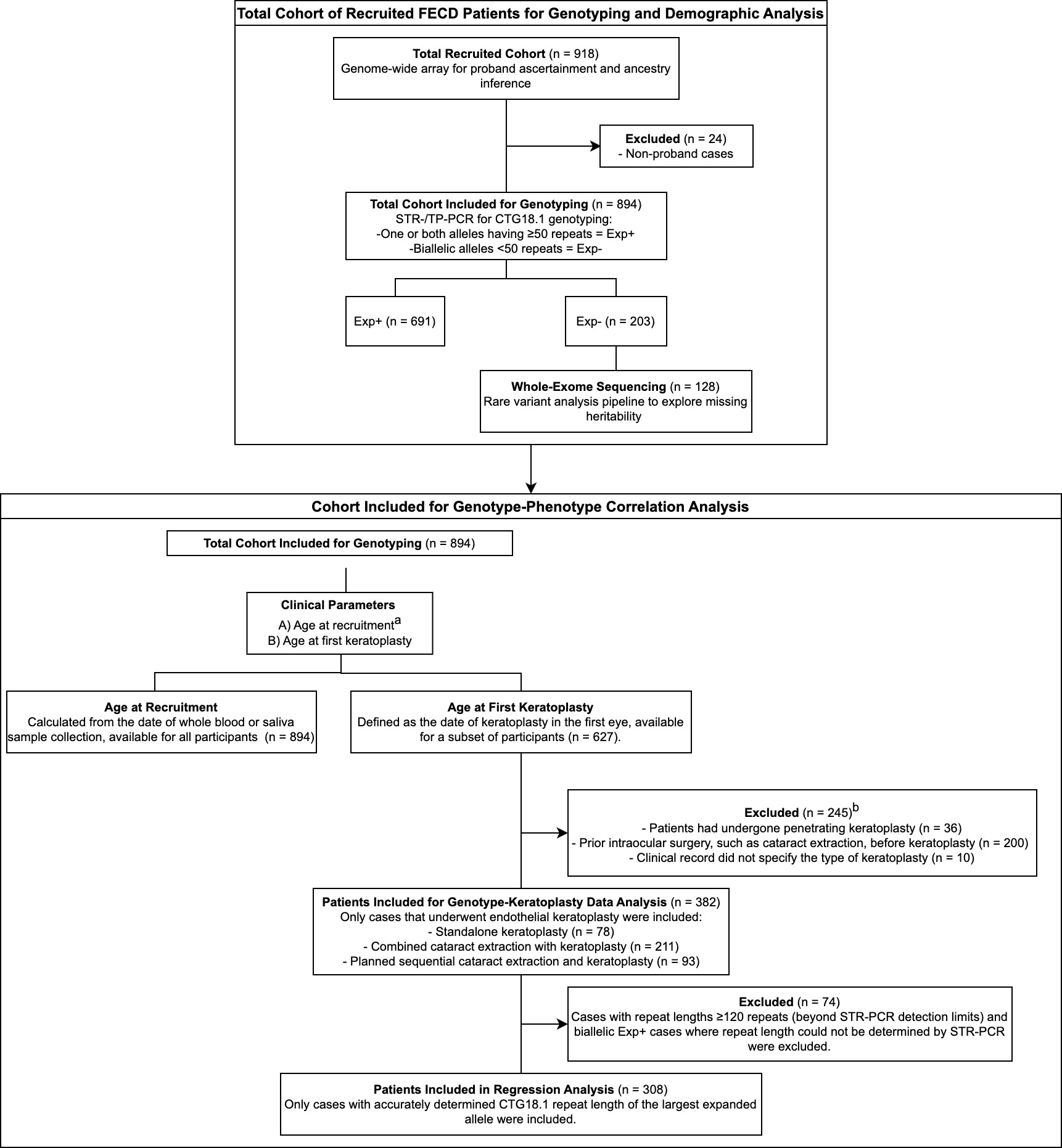
