## Supplementary material for "Genetic and Demographic Determinants of Fuchs Endothelial Corneal Dystrophy Risk and Severity": eTable 1

**Online-Only Tables**

|  | **Number of cases** | **CTG18.1 Exp+** | | | **CTG18.1 Exp-** |
| --- | --- | --- | --- | --- | --- |
|  |  | **Cases with ≥1 expanded allele** | **Monoallelic expanded cases** | **Biallelic expanded cases** |  |
| Cases Meeting Inclusion/Exclusion Criteria (N=649/894)^a^ | | | | | |
| **Number of patients operated** | 382/649 (58.9%) | 323/480 (67.3%) | 300/446 (67.3%) | 23/34 (67.6%) | 59/169 (34.9%) |
| Female | 228/411 (55.5%) | 181/283 (64.0%) | 170/266 (63.9%) | 11/17 (64.7%) | 47/128 (36.7%) |
| Male | 154/238 (64.7%) | 142/197 (72.1%) | 130/180 (72.2%) | 12/17 (70.6%) | 12/41 (29.3%) |

**eTable 1. Keratoplasty data of Fuchs endothelial corneal dystrophy (FECD) patient cohort stratified by sex and CTG18.1 genotype.** Exp+, CTG18.1 expansion positive allele defined as ≥50 CTG repeats; Exp-, CTG18.1 expansion positive allele defined as <50 CTG repeats; MEH, Moorfields Eye Hospital; GUH, General University Hospital, IQR, interquartile range. ^a^Inclusion criteria: endothelial keratoplasty, whether as a standalone procedure, combined with phacoemulsification, or sequentially planned after phacoemulsification; Exclusion criteria: Penetrating keratoplasty, prior intraocular surgery, and unspecified keratoplasty type; the denominators include the total cohort, encompassing both operated and unoperated cases, after excluding those that meet the exclusion criteria.
