## Supplementary material for "Genetic and Demographic Determinants of Fuchs Endothelial Corneal Dystrophy Risk and Severity": eTable 2

|  | **Number of Individuals** | **CTG18.1 Exp-** | **CTG18.1 Exp+** | | | **Biallelic:monoallelic ratio** | **Expected biallelic expanded cases^c^** |
| --- | --- | --- | --- | --- | --- | --- | --- |
|  |  |  | **Cases with ≥1 expanded allele** | **Monoallelic expanded cases** | **Biallelic expanded cases** |  |  |
| FECD cases of European Ancestry | 829 | 160 | 669 | 623 | 46 | 1:14 | 7 |
| Ethnicity-matched Controls^a^ | 550 | 527 | 23 | 23 | 0 | 1:94^b^ | <1 |

**eTable 2. Summary of CTG18.1 expansion status within Fuchs endothelial corneal dystrophy (FECD) probands of European ancestry and ethnicity-matched controls, where the derived allele frequency was used for expected and observed homozygous:heterzygous ratio calculation.** Exp+, CTG18.1 expansion positive allele defined as ≥50 CTG repeats; Exp-, CTG18.1 expansion positive allele defined as <50 CTG repeats. ^a^Data presented from Zarouchlioti et al. 2018.^1^ ^b^Control ratio under Hardy-Weinberg equilibrium that was confirmed in the control group (P=.617). ^c^Expected number of biallelic expanded cases based on control biallelic:monoallelic ratio and observed monoallelic expanded cases.
