## Supplementary material for "Genetic and Demographic Determinants of Fuchs Endothelial Corneal Dystrophy Risk and Severity": eTable 3

| **Model – single longest CTG18.1 allele measured per individual in all Exp+ patients** | **Age at first keratoplasty** | | | | |
| --- | --- | --- | --- | --- | --- |
| **Variables** | **Number of cases** | **Adjusted R^2^** | **Regression coefficient** | **95% CI** | ***P* value** |
| CTG18.1 repeat length of the largest allele | 308 | 0.013 | -0.087 | -0.162 to -0.012 | 0.024 |

**eTable 3. Linear regression models analyzing the relationship between CTG18.1 repeat length of the largest allele and age at first keratoplasty.** Exp+, CTG18.1 expansion positive allele defined as ≥50 CTG repeats.
