## Supplementary material for "Genetic and Demographic Determinants of Fuchs Endothelial Corneal Dystrophy Risk and Severity": eTable 4

| **Gene**  **CEC TPM** | **Transcript**  **(ENST00000-)** | **Patient ID** | **Variant** | **CDS** | **Protein** | **CADD** | **gnomAD (v3.1.2)** | | **Age at first keratoplasty (Years)** | **Sex** | **Ancestry^a^** | **Family history** | **Site** | **Previously reported** |
| --- | --- | --- | --- | --- | --- | --- | --- | --- | --- | --- | --- | --- | --- | --- |
|  |  |  |  |  |  |  | **Total** | **Max (Population)** |  |  |  |  |  |  |
| *AGBL1*  TPM = 0 | 614907.3 | P824 | chr15-86247772-A-T | c.628A>T | p.(Ile210Phe) | 18.3 | - | - | 66-70 | F | EUR | Unknown | MEH | No |
|  |  | P371 | chr15-86295271-T-C | c.2237T>C | p.(Leu746Pro) | 25.2 | - | - | 70-75 | F | EUR | Unknown | MEH | No |
|  |  | P864 | chr15-86674435-C-T | c.3157C>T | p.(Arg1053Trp) | 36 | 0.001952  297/152158 | 0.008353 (ASJ)  29/3472 | NS | M | EUR | Unknown | MEH | No |
|  |  | P759 | chr15-86907208-C-T | c.3280C>T | p.(Arg1094Trp) | 16.8 | 0.001007  153/152002 | 0.003020 (AFR)  125/41394 | 40-45 | F | AFR | Unknown | MEH | No |
| *LOXHD1*  TPM = 0.02 | 642948.1 | P2185 | chr18-46610832-T-G | c.703A>C | p.(Lys235Gln) | 26.0 | 0.00000656  1/152220 | 0.0000147  1/68030 | NS | M | EUR | No | GUH | No |
|  |  | P327 | chr18-46592017-G-A | c.1570C>T | p.(Arg524Cys) | 30 | 0.002833  431/152158 | 0.008903 (AMR)  136/15276 | 66-70 | F | EUR | Unknown | MEH | No |
|  |  | P354 | chr18-46592017-G-A | c.1570C>T | p.(Arg524Cys) | 30 | 0.002833  431/152158 | 0.008903 (AMR)  136/15276 | 70-75 | F | EUR | Unknown | MEH | No |
|  |  | P722 | chr18-46541815-G-A | c.3874C>T | p.(Leu1292Phe) | 25.7 | 0.0003088  47/152222 | 0.001223 (FIN)  13/10626 | 70-74 | M | EUR | Unknown | MEH | No |
|  |  | P2180 | chr18-46541787-A-C | c.3902T>G | p.(Leu1301Arg) | 16.2 | 0.00000657  1/152206 | 0.00006542 (AMR)  1/15286 | NS | F | EUR | No | GUH | No |
|  |  | P759 | chr18-46533293-C-T | c.4244G>A | p.(Arg1415Gln) | 27.3 | 0.0006768  103/152190 | 0.002293 (AFR)  95/41436 | 40-45 | F | AFR | Unknown | MEH | No |
|  |  | P523 | chr18-46529227-G-A | c.4480C>T | p.(Arg1494Ter) | 42 | 0.0006249  95/152026 | 0.001152 (ASJ)  4/3472 | 70-75 | F | EUR | Unknown | MEH | No |
|  |  | P399 | chr18-46522163-G-A | c.5023C>T | p.(Arg1675Cys) | 19.2 | 0.001512  230/152102 | 0.006743 (AMR)  103/15276 | 56-60 | F | AFR | Unknown | MEH | No |
|  |  | P788 | chr18-46505914-G-T | c.5802C>A | p.(Asn1934Lys) | 21.3 | 0.003082  469/152198 | 0.005624 (AMR)  86/15292 | 76-80 | F | EUR | Unknown | MEH | No |
|  |  | P522 | chr18-46477568-G-A | c.6726C>T | p.(Thr2242Thr) | 17.6 | 0.00002627  4/152258 | 0.00004823 (AFR)  2/41472 | 66-70 | F | EUR | Unknown | MEH | No |
|  |  | P737 | chr18-46477553-G-A | c.6741C>T | p.(Ala2247Ala) | 18.9 | 0.003061  466/152228 | 0.00463 (NFE)  315/68032 | 76-80 | F | NA | Unknown | MEH | No |

**eTable 4. Summary of rare variants identified in *LOXHD1* and *AGBL1* from 128 FECD CTG18.1 Exp- probands analysed by exome sequencing.** The identifiers used in this study do not disclose the identity of any participants beyond the immediate research team. CEC, corneal endothelial cells; TPM, transcript per million; CADD, Combined Annotation Dependent Depletion; MAF, minor allele frequency; EUR, European; AFR, African American/African; FIN, Finnish; NFE, non-Finnish European, ASJ, Ashkenazi Jews; M, male; F, female; NS, no surgery; MEH, Moorfields Eye Hospital London; GUH, General University Hospital Prague.  ^a^FRAPOSA predicted ancestry
